# Pulmonary Embolism Obstruction Model to Evaluate Clinical Deterioration

**DOI:** 10.1101/2023.07.27.23293295

**Authors:** Jian-Kuan Yeh, Po-Wei Chen, Wei-Ting Chang, Pin-Hsuan Chiu, Pei-Fang Su, Chih-Hsin Hsu, Chih-Chan Lin, Hsien-Yuan Chang

## Abstract

Hemodynamic instability may develop in patients with acute pulmonary embolism (PE) days after the emboli event. Simplified methods to predict clinical deterioration are currently lacking. This retrospective cohort study included patients diagnosed with acute. The aim is to develop a simplified imaging model with good clinical accessibility to predict the clinical deterioration of patients with acute PE. This study included patients with acute PE under the International Classification of Disease, ninth or tenth revision. Seven models based on computed tomography pulmonary angiography (CTPA) were developed based on the location (central versus peripheral) and the degree (nearly total versus partial) of obstruction. The outcome includes clinical deterioration, which is defined as death from PE, cardiopulmonary resuscitation, mechanical ventilation, vasopressor therapy, thrombolysis, catheter-directed therapy, and surgical embolectomy. Logistic regression analysis was used to test the association between different models and clinical deterioration. The area under the receiver operating characteristic curve (AUC) was used to test the predictive ability. The category-free net reclassification improvement (NRI) and integrated discrimination improvement (IDI) were used to quantify the improvement of the proposed models plus the simplified Pulmonary Severity Index (sPESI) compared with the sPESI alone. Calculating the nearly totally obstructed 20 peripheral arteries provides good predictive ability in the seven models (AUC: 0.77). Calculating nearly totally obstructed 20 peripheral segments can predict clinical deterioration. Obstruction on CTPA combined with the sPESI increased the ability to predict clinical deterioration compared to the sPESI alone and may be used as guidance in clinical decision-making.

**Clinical perspective:** A simple model based on computed tomography pulmonary angiography (CTPA) to predict clinical deterioration in patients with acute pulmonary embolism (PE) is currently lacking.

This retrospective study included 210 patients and used the model for calculating the nearly totally obstructed segmental pulmonary arteries as an efficient and simple method to predict clinical deterioration. This model added to the simplified PE severity index (sPESI) has an increased predictive ability compared to the sPESI alone. CTPA images can predict the clinical deterioration of patients with acute PE and may assist in clinical decision-making.

## Introduction

Acute pulmonary embolism (PE), following myocardial infarction and stroke, is the third most common cardiovascular disease. ^1^ The annual incidence of PE is approximately 1 per 1,000 people. ^2^ PE occurs when emboli occlude the pulmonary arteries and may cause complications, such as arrhythmias, right ventricular (RV) failure, cardiogenic shock, and even death. ^3^ Effective risk stratification is crucial for treating PE, as it helps identify patients that may benefit from thrombolytic therapies on top of systemic anticoagulation. The 2019 European Society of Cardiology (ESC) guideline on PE classifies patients into different risk groups based on the presence of hemodynamic instability, RV dysfunction, elevated cardiac enzymes, and the PE risk index (PESI). ^4^ Thrombolytic therapy is recommended for high-risk patients with PE, who are defined as having hemodynamic instability. The benefit of early reperfusion in patients without hemodynamic instability but with other clinical indicators of poor outcomes remains unknown. However, patients with acute PE may develop acute RV failure days after the emboli event, followed by hemodynamic instability and clinical deterioration. Prompt recognition of patients who are prone to clinical deterioration is important, but clinical predictors are currently lacking.

Miller et al. first developed an index based on invasive pulmonary angiography to describe the thrombus burden and the severity of PE to evaluate the treatment response of systemic thrombolysis. However, the clinical utility of the Miller index is limited by its invasive nature. ^5^ Qanadli et al. later developed an index (PAOI) based on computed tomography pulmonary angiography (CTPA) which correlated well with the Miller index and could predict RV dilatation. The Qanadli index is of limited clinical utility due to its complexity in score calculation. ^6^ Studies revealed that the Qanadli index is strongly predictive of high-risk patients. ^7,8^ However, other studies revealed no significant correlation between the thrombus burden and clinical risks. ^9,10^

This study primarily aimed to develop a simplified imaging model with good clinical accessibility to predict the clinical deterioration of patients with acute PE.

## Materials and methods

### Materials

This retrospective study used the cardiovascular databank of the National Cheng Kung University Hospital. The enrollment period spanned from January 1, 2008, to December 31, 2019, and included patients diagnosed with PE under the International Classification of Disease, ninth or tenth revision. This study adhered to the Declaration of Helsinki and obtained approval from the Human Research and Ethics Committee of the National Cheng Kung University Hospital (IRB number: B-ER-109-102). An additional group of patients from Chi Mei Medical Center, which is another tertiary medical center, from January 1, 2008, to December 31, 2021, was included for external validation. The external validation group also received approval from the Human Research and Ethics Committee of the Chi Mei Medical Center (IRB number: CMMC11011-002).

This study applied certain exclusion criteria. Patients with septic, tumor, or fat emboli, tumor invasion or pulmonary artery encasement, and stump thrombosis were excluded from the study, as well as patients with only segmental or subsegmental PE, where the thrombi were too small to cause clinical symptoms. Further, cases with poor image quality or no diagnostic CTPA were excluded. Furthermore, patients with CT-defined chronic thrombus were excluded due to differing prognoses. The simplified PESI (sPESI) was calculated. ^11^ The administration of systemic or catheter-directed thrombolysis was recorded for each patient.

### Obstruction index determination

Two specialists, who were unaware of the patient’s clinical data, evaluated the images. Two indices, namely the Miller obstruction index and the Qanadli index, were calculated based on CTPA. The Miller obstruction index is calculated as **Σ** (*n.d*), where n represents the number of segmental arteries with thrombus (range: 1–16) and d is the presence (1) or absence (0) of obstruction. According to Miller et al., the right pulmonary artery has nine major segmental arteries (three to the upper lobe, two to the middle lobe, and four to the lower lobe), and the left pulmonary artery has seven major segmental arteries (two to the upper lobe, two to the lingula, and four to the lower lobe). ^5^ The Qanadli index is calculated as **Σ** (*n.d*)/40 × 100, where n is the number of segmental arteries with thrombus (ranging from 1 to 20), and d is the degree of obstruction (range: 0–2). Both pulmonary arteries have ten segmental arteries (three to the upper lobes, three to the middle lobe and lingula, and four to the lower lobes). The degree of obstruction ranges from 0 (no obstruction) to 1 (partial obstruction) to 2 (nearly total obstruction). ^6^

The degree of obstruction of the central segments was also scored, using the same scale of 0 (no obstruction) to 1 (partial obstruction) to 2 (nearly total obstruction), to comprehensively assess the pulmonary circulation and evaluate the entire pulmonary vasculature. The central pulmonary artery was divided into seven segments based on the lung lobes they supply: the main pulmonary artery, the proximal, middle, and distal parts of bilateral pulmonary arteries. The proximal segment of the right and left pulmonary arteries represents the area between the branching from the main pulmonary artery to the branching of the segmental arteries to the upper lobe. The middle segment lies between the upper lobe segmental arteries and the middle lobe segmental arteries, while the distal segment is distal to the segmental arteries to the middle lobe. The transverse section of the pulmonary artery primarily determines the obstruction percentage (Figure 1).

**Figure 1.**
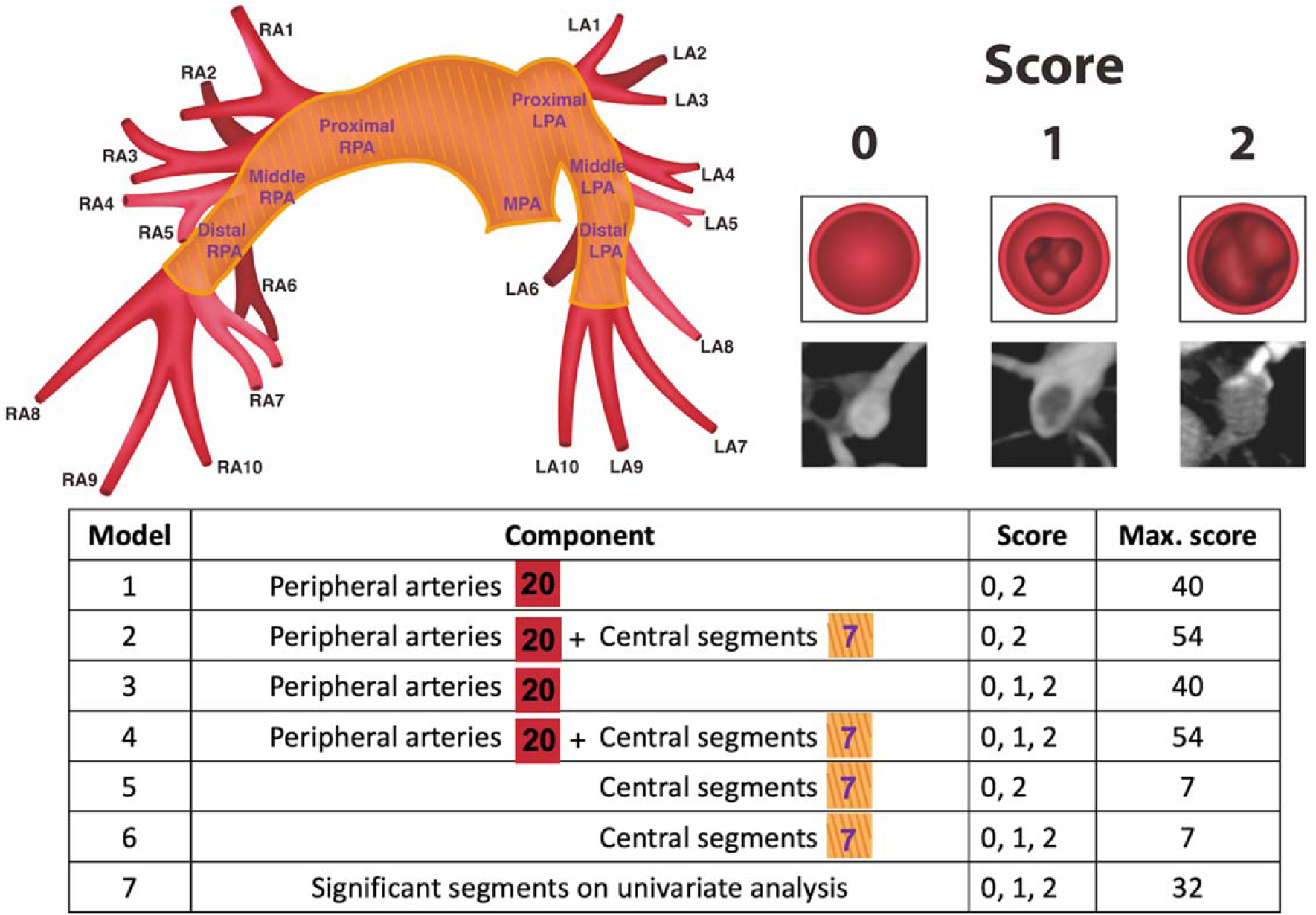
Illustration of Models based on Central and Peripheral Pulmonary Arteries and Degree of Obstruction. The left upward illustration is a schematic representation of the pulmonary arterial vasculature. The area in red represents the 20 peripheral segmental pulmonary arteries and the area shaded in yellow represents the central pulmonary arteries, which is further divided into the proximal, middle, and distal parts based on the lung lobes which they supply. The right upward illustration is the scores based on levels of obstruction. The illustrations and the reference on computed tomography are listed below. A score of 0 indicates no thrombus, a score of 1 indicates partial obstruction, and a score of 2 indicates nearly total obstruction. The below table is an explanation of models 1 to 7. Model 1 represents the sum of nearly total obstructed peripheral arteries. Model 2 is the sum of nearly total obstructed central and peripheral arteries. Model 3 includes the sum of peripheral pulmonary artery obstruction, which is the Qanadli index. Model 4 combines central and peripheral pulmonary artery obstruction. Model 5 comprises the sum of nearly total obstructed central arteries. Model 6 is the sum of central pulmonary artery obstruction. Abbreviations: LA1-10, 1^st^ to 10^th^ segmental pulmonary arteries of the left pulmonary artery; LPA, left pulmonary artery; RA1-10, 1^st^ to 10^th^ segmental pulmonary arteries of the right pulmonary artery; RPA, right pulmonary artery.

### Proposed models

Six models were developed based on the location (central versus peripheral) and the degree (nearly total versus partial) of obstruction. The peripheral arteries are the segmental pulmonary arteries. Model 1 represents the sum of nearly totally obstructed peripheral arteries. Model 2 is the sum of nearly totally obstructed central and peripheral arteries. Model 3 contains the Qanadli index, which is the sum of peripheral pulmonary artery obstruction. Model 4 combines central and peripheral pulmonary artery obstruction. Model 5 comprises the sum of nearly totally obstructed central arteries. Model 6 is the sum of central pulmonary artery obstruction. A seventh model, in addition to the six pre-specified models, was developed by performing univariate logistic regression and selecting the statistically significant parts of pulmonary arteries that impact the outcome (Figure 1). The analysis included the combination of models 1 to 7 together with the sPESI. Further discussions were conducted when there is a discrepancy in judgment until a conclusion is reached.

### Outcomes

Two specialists carefully reviewed the electric medical records of enrolled patients. The outcome included clinical deterioration, which is death from PE, cardiopulmonary resuscitation, mechanical ventilation, vasopressor therapy for systemic arterial hypotension, thrombolysis, catheter-directed therapy, and surgical embolectomy^12^. Each inpatient was followed up until discharged, while outpatients were followed up for 1 month. PE-related death was analyzed as a separate outcome.

### Statistical analysis

Continuous data are presented as the mean ± standard deviation while dichotomous data as numbers and percentages. The Wilcoxon Rank Sum test was used for comparisons of continuous variables. Fisher’s exact test was used for categorical variables.

Variables associated with clinical deterioration were identified using univariate logistic regression analysis along with Bonferroni correction and false discovery rate. Multivariate logistic regression was used to analyze the combination of different models plus sPESI. Model performance was calculated by the area under the curve (AUC) of the receiver operating characteristic curve. The accuracy, sensitivity, and specificity were calculated. The category-free NRI and IDI were used to quantify the improvement of the proposed models plus sPESI in comparison with the reference model, which is the sPESI in our study. The Bland-Altman analysis of agreement and the interanalysis correlation coefficient were used to assess the intra- and inter-rater reliability. Finally, a nomogram combining the imaging model and sPESI was proposed. Statistical software R (Version 4.0.2 for Windows) was used for all statistical tests. All statistical tests were 2-sided, and a *P*-value of <0.05 was considered statistically significant.

## Results

### Study population

A total of 509 patients with PE were retrospectively screened. The exclusions comprised 111, 37, 40, and 111 patients with only segmental or subsegmental PE, chronic PE, other PE etiologies, and suboptimal image quality for evaluation, respectively (Figure S1). Finally, the analysis included 210 patients (age: 65 ± 16 years; male: 40%), including 170 (81%) with no clinical deterioration and 40 (19%) with clinical deterioration. The baseline clinical characteristics were balanced between patients with and without clinical deterioration apart from the sPESI. The sPESI was statistically significantly higher in the group with clinical deterioration (2.55 ± 1.47 vs. 1.42 ± 1.13, *P* < 0.01). Among the 40 patients with clinical deterioration, 21 (53%) received systemic thrombolysis. The Miller index and the Qanadli index were higher in the group with clinical deterioration (Table 1).

**Table 1.** Baseline Characteristics of Patients with Acute Pulmonary Embolism with and without Clinical Deterioration.

|  | Overall (N = 210) | With clinical deterioration (N = 40) | Without clinical deterioration (N = 170) | p-value |
| --- | --- | --- | --- | --- |
| Age (years old) | 64.98 ± 16.18 | 65.47 ± 17.44 | 64.86 ± 15.92 | .66 |
| Sex (male) | 85 (40%) | 18 (45%) | 67 (39%) | .59 |
| Body mass index (kg/m<sup>2</sup>) | 25.99 ± 5.84 | 25.26 ± 4.93 | 26.16 ± 6.03 | .98 |
| sPESI | 1.63 ± 1.28 | 2.55 ± 1.47 | 1.42 ± 1.13 | <.01 |
| Cancer | 65 (31%) | 12 (30%) | 53 (31%) | .99 |
| Hypertension | 115 (55%) | 23 (58%) | 92 (54%) | .73 |
| Diabetes mellitus | 43 (20%) | 6 (15%) | 37 (22%) | .39 |
| Coronary artery disease | 19 (9%) | 3 (8%) | 16 (9%) | .99 |
| Heart failure | 27 (13%) | 6 (15%) | 21 (13%) | .61 |
| Atrial fibrillation | 26 (12%) | 7 (18%) | 19 (11%) | .29 |
| Old stroke | 19 (9%) | 3 (8%) | 16 (9%) | .99 |
| Risk |  |  |  |  |
| High | 15 (7%) | 11 (28%) | 4 (2%) |  |
| Intermediate | 119 (57%) | 23 (58%) | 96 (56%) | <.01 |
| Low | 76 (35%) | 6 (15%) | 70 (41%) |  |
| Systemic t-PA | 21 (10%) | 21 (53%) | 0 (0%) | <.01 |
| Miller index | 11.30 ± 4.33 | 12.88 ± 3.28 | 10.93 ± 4.47 | .01 |
| Qanadli index | 20.37 ± 9.31 | 25.27 ± 7.90 | 19.21 ± 9.26 | <.01 |

Data are presented as mean ± SD or N (%)
Abbreviations: sPESI, simplified Pulmonary Embolism Severity Index.

This study retrospectively screened 319 patients from another tertiary medical center, with 209 patients being excluded (Figure S2). Finally, the external validation included 109 patients (age: 64 ± 17 years; male: 43%), including 84 (77%) with no clinical deterioration and 25 (23%) with clinical deterioration (Table S1).

### Degree of obstruction and clinical outcomes

The sPESI demonstrated predictive ability for mortality in patients with acute PE, with an AUC of 0.740. However, both the Miller and Qanadli indices had limited predictive ability of mortality, with AUC values of 0.635 and 0.578, respectively. Additionally, the sPESI exhibits the best performance in predicting clinical deterioration, with an AUC of 0.724. The Miller and Qanadli indices can predict clinical deterioration, with AUC values of 0.624 and 0.678, respectively (Figure 2). The external validation group demonstrated similar results (Figure S3). The sPESI demonstrated predictive ability for both mortality and clinical deterioration, while the Miller and Qanadli indices have low predictive ability for mortality.

**Figure 2.**
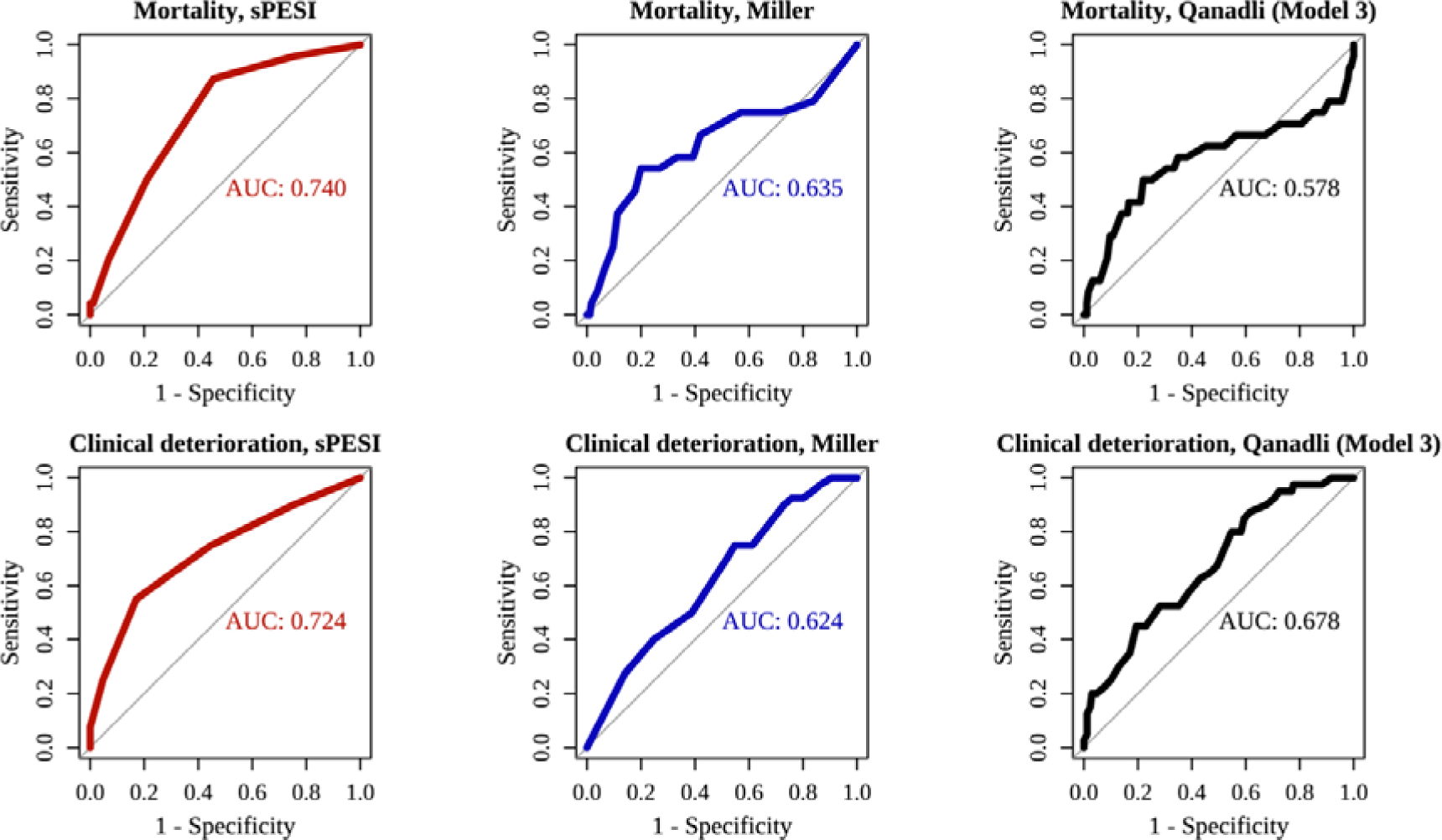
The Area Under the Receiver Operating Characteristic for the sPESI, Miller Index, and Qanadli Index to Predict Mortality and Clinical Deterioration. Abbreviations: sPESI, simplified Pulmonary Embolism Severity Index

### Comparison of the predictive ability of different models

We compared models 1 to 6 to assess the impact of different locations (central versus peripheral) and levels of obstruction (partial versus nearly total) on clinical deterioration. Models 1 and 2 were compared to examine whether adding nearly total occluded central pulmonary arteries will improve the predictive ability. Models 1 and 2 exhibit nearly identical predictive abilities with an AUC of 0.70, along with the same accuracy of 0.82. Furthermore, models 3 and 4 were compared to explore the additive effect of central pulmonary arteries. Model 4 demonstrated similar predictive ability (AUC: 0.69) and accuracy (0.81) compared to model 3 (AUC: 0.68 and accuracy: 0.81). Models 5 and 6 were designed to analyze the predictive ability of obstruction of central pulmonary arteries alone. Model 6 has a better predictive ability (AUC: 0.68) compared to model 5 (AUC: 0.63), while both models have similar accuracy (0.81). Finally, model 7 is the summation of statistically significant variables calculated on univariate analysis. It is composed of the first, third, sixth, and eighth segmental arteries of the right pulmonary artery, the third to ninth segmental arteries of the left pulmonary artery, the middle right pulmonary artery, the main pulmonary artery, and the left proximal to distal pulmonary arteries (Table S2). Model 7 demonstrated the best predictive ability among all the models (AUC: 0.72) (Table 2).

**Table 2.** The ROC Analysis of Different Models with and without sPESI in Predicting Clinical.

| Models | Peripheral | Central | Partial<br>occlusion | Nearly<br>total<br>occlusion | sPESI | AUC (95% CI) | Accuracy |
| --- | --- | --- | --- | --- | --- | --- | --- |
| Model 1 | ● |  |  | ● |  | 0.6996 (0.6148-0.7843) | 0.8238 |
| Model 2 | ● | ● |  | ● |  | 0.6996 (0.6160-0.7832) | 0.8238 |
| Model 3 | ● |  | ● | ● |  | 0.6777 (0.5886-0.7669) | 0.8095 |
| Model 4 | ● | ● | ● | ● |  | 0.6895 (0.6015-0.7774) | 0.8143 |
| Model 5 |  | ● |  | ● |  | 0.6254 (0.5342-0.7167) | 0.8095 |
| Model 6 |  | ● | ● | ● |  | 0.6802 (0.5915-0.7690) | 0.8095 |
| Model 7 | Significant segments on univariate analysis |  |  |  |  | 0.7222 (0.6404-0.8040) | 0.819 |
| Model 1 + sPESI | ● |  |  | ● | ● | 0.7753 (0.6914-0.8592) | 0.8619 |
| Model 2 + sPESI | ● | ● |  | ● | ● | 0.7765 (0.6914-0.8615) | 0.8571 |
| Model 3 + sPESI | ● |  | ● | ● | ● | 0.7777 (0.6937-0.8617) | 0.8619 |
| Model 4 + sPESI | ● | ● | ● | ● | ● | 0.7824 (0.6990-0.8657) | 0.8667 |
| Model 5 + sPESI |  | ● |  | ● | ● | 0.7418 (0.6467-0.8370) | 0.8333 |
| Model 6 + sPESI |  | ● | ● | ● | ● | 0.7659 (0.6752-0.8566) | 0.8476 |
| Model 7 + sPESI | Significant segments on univariate analysis |  |  |  | ● | 0.8004 (0.7204-0.8805) | 0.8619 |

Abbreviations: AUC, area under curve; ROC, receiver operating characteristic; sPESI,
simplified Pulmonary Embolism Severity Index

The external validation group analysis revealed similar results. Model 6, which is the summation of partially and totally occluded central pulmonary arteries, exhibited the best predictive ability in the external validation group (Table S3).

### Integrated discrimination index (IDI) and net reclassification index (NRI)

The addition of sPESI to models 1 to 7 resulted in an improved predicting value, with an increased mean AUC from 0.68 to 0.77 and increased mean accuracy from 0.82 to 0.86. Model 7 plus sPESI demonstrated the best predictive ability with an AUC of 0.80 (95% confidence interval [CI]: 0.72–0.88), while model 4 plus sPESI achieved the highest accuracy of 0.87 (Table 2, Table S4). The external validation group analysis revealed similar results, with an increased mean AUC from 0.73 to 0.84 and increased mean accuracy from 0.79 to 0.83 after adding sPESI to the models (Table S3). Statistically significant improvements in predicting clinical deterioration were observed based on IDI, with the IDI ranging from 0.02 to 0.09, when comparing models 1 to 7 added to the sPESI with sPESI alone. The NRI demonstrated an increased predictive ability of models 1–7 with the addition of sPESI (Table 3). The IDI and NRI of the external validation group revealed an improvement in the predicting ability of the models in combination with sPESI compared to sPESI alone (Table S5). Overall, the combination of clinical risk factors and the obstruction severity on CTPA provides a better prediction of clinical deterioration.

**Table 3.**
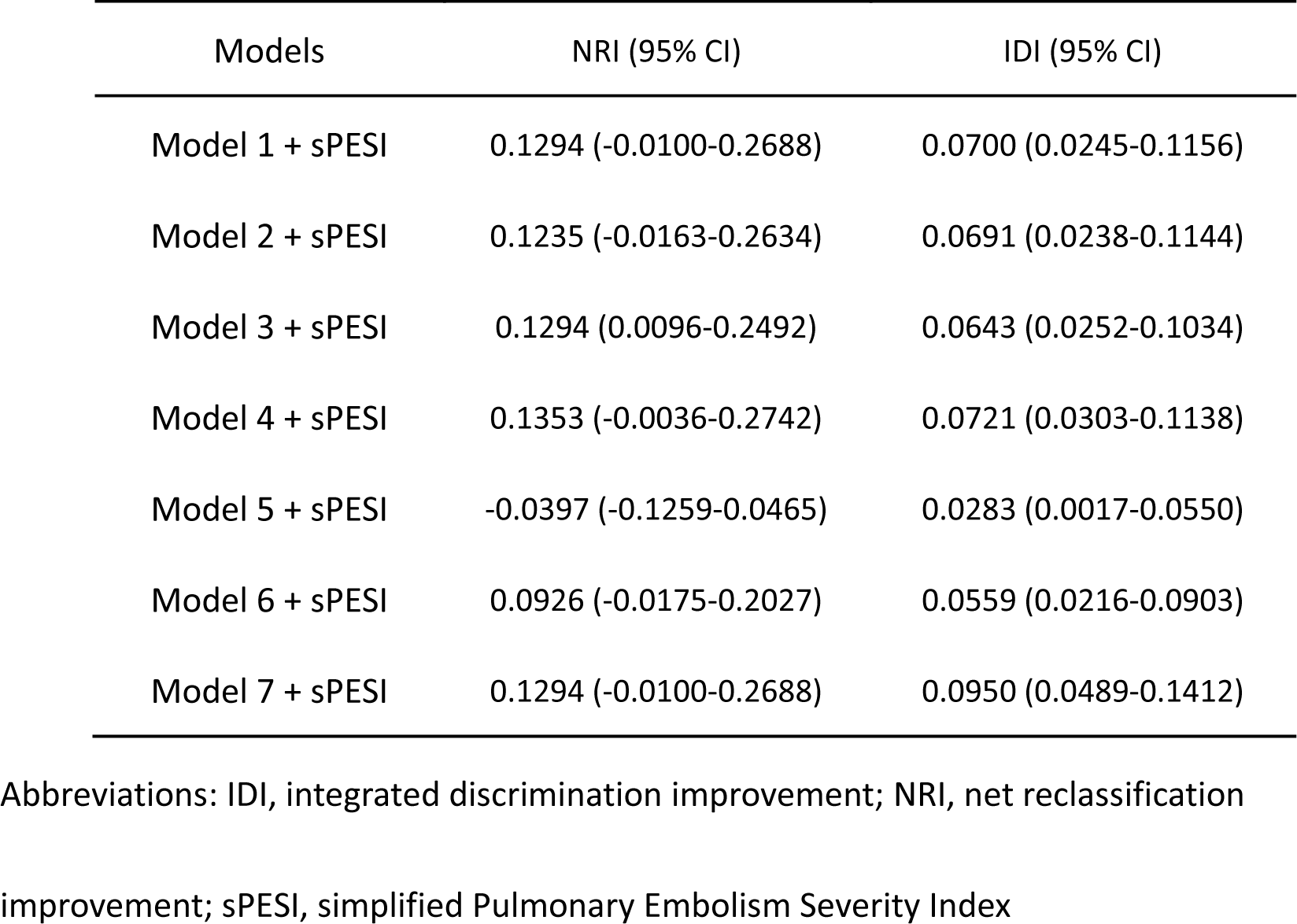
The NRI and IDI Comparing Models Plus sPESI with sPESI.

| Models | NRI (95% CI) | IDI (95% CI) |
| --- | --- | --- |
| Model 1 + sPESI | 0.1294 (-0.0100-0.2688) | 0.0700 (0.0245-0.1156) |
| Model 2 + sPESI | 0.1235 (-0.0163-0.2634) | 0.0691 (0.0238-0.1144) |
| Model 3 + sPESI | 0.1294 (0.0096-0.2492) | 0.0643 (0.0252-0.1034) |
| Model 4 + sPESI | 0.1353 (-0.0036-0.2742) | 0.0721 (0.0303-0.1138) |
| Model 5 + sPESI | -0.0397 (-0.1259-0.0465) | 0.0283 (0.0017-0.0550) |
| Model 6 + sPESI | 0.0926 (-0.0175-0.2027) | 0.0559 (0.0216-0.0903) |
| Model 7 + sPESI | 0.1294 (-0.0100-0.2688) | 0.0950 (0.0489-0.1412) |

Abbreviations: IDI, integrated discrimination improvement; NRI, net reclassification
improvement; sPESI, simplified Pulmonary Embolism Severity Index

Nomograms based on the models plus sPESI can be used as a tool in the clinical setting to predict prognosis (Figure S4).

### Inter- and intra-rater variability

Model 3, which is the Qanadli index, is used to test the intra- and inter-rater variability. The intra- and inter-rater correlation coefficients of model 3 were 0.995 (95% CI: 0.993– 0.996) and 0.989 (95% CI: 0.986–0.992), respectively (Table S6). The Bland-Altman analysis revealed no intra-rater and inter-rater bias in the calculation of model 3 scores (Figure S5).

## Discussion

Our study revealed that the sPESI can predict clinical deterioration in patients with PE, and the obstruction level on CTPA can predict mortality and clinical deterioration. After the addition of central segments and partially occluded segments, the predictivity ability is nearly identical. Calculating the nearly totally obstructed 20 peripheral segments is an efficient method that balances predictive ability and clinical usability. Combining the severity of obstruction on the image with sPESI improves the predictive ability for clinical deterioration compared to sPESI alone.

Animal studies have revealed that acute pulmonary artery obstruction increases pulmonary artery pressure and pulmonary vascular resistance. ^13,14^ Severe pulmonary hypertension may cause RV failure, clinical deterioration, and hemodynamic instability. Currently, discrepancies exist in the literature regarding whether PA clot burden could reflect severity. Some studies revealed thrombus burden as a significant predictor of death in patients with acute PE^15–17^, but others have not. ^9,10^ The ESC recommended against using the anatomical burden and emboli characteristics to determine PE severity. ^18^ Our study revealed a positive relationship between the clot burden and both mortality and clinical deterioration, indicating the total burden as not the only affecting factor of clinical deterioration. The obstruction site together with the clot burden is influential in predicting clinical deterioration.

Calculating only the occluded central pulmonary arteries had the lowest predictability of clinical deterioration, possibly because proximal artery occlusion does not necessarily reduce distal blood flow. Including the clot burden of the central pulmonary arteries to the peripheral pulmonary arteries yielded similar predictive ability compared to calculating obstructed peripheral pulmonary arteries alone. Similarly, including partially occluded pulmonary arteries yielded similar predictive ability as to calculating the nearly total occluded arteries alone. This indicates a forward blood flow passing through partially occluded arteries to the distal circulation.

The 2019 ESC guidelines on PE management classify patients into high, intermediate, and low-risk groups, and recommended treatment options based on risk strata. However, approximately 5% of patients initially classified as intermediate risk will develop hemodynamic decompensation. ^19^ Our study aims to identify methods to predict clinical deterioration in this specific group of patients, where the benefits of early thrombolytic therapy may outweigh the risk of bleeding. Huang et al. used a three-dimensional CT method to estimate the total embolic burden and revealed a positive correlation between the total embolic volume and impending shock. ^20^ Additionally, our models demonstrated a positive correlation between the severity of obstruction and clinical deterioration, offering a straightforward and easily applicable method to determine severity in the clinical setting.

The sPESI, which includes age, cancer, chronic cardiopulmonary diseases, heart rate, systolic blood pressure, and oxyhemoglobin levels, can predict the 30-day mortality of patients with PE. ^11^ Our study revealed that the sPESI predicted clinical deterioration. However, adding the extent of obstruction of pulmonary improves the ability to predict clinical deterioration since the variables included in the sPESI are not specific to PE alone. Our models, increased the predictive ability when added to sPESI, as evidenced by positive NRI and IDI values. The low inter- and intra-observer variability indicates good agreement between the two investigators. The nomogram combining model 1 (totally obstructed peripheral arteries) and the sPESI (Figure S4) is a simple and reproducible tool to aid physicians in assessing the prognosis of patients with acute PE.

Current treatment guidelines for acute PE recommend systemic thrombolysis for patients who present with hemodynamic instability, ^21,22^ but the management of patients without hemodynamic instability but with unstable clinical features or evidence of RV dysfunction on imaging remains uncertain. Previous studies revealed that treating patients with acute PE with systemic thrombolysis on a background of heparin lowers the risk of developing in-hospital death, clinical deterioration, or hemodynamic decompensation. ^19,23^ Additionally, a meta-analysis demonstrated that treating acute PE with thrombolytic therapy lowers mortality, but increases the risk of major bleeding. ^24^ The benefit of thrombolytic therapy may outweigh the risk of bleeding in certain normotensive patients with acute PE with high-risk features. Therefore, developing a more precise prediction method to identify the patients prone to clinical deterioration is essential. Our study may help identify the patients suitable for further investigation into whether the benefit of thrombolytic therapy may outweigh the risk of bleeding.

This study has several limitations, including its small sample size and its retrospective design, which prevents assessment of the effects on clinical decision-making and patient outcomes. Additionally, the inclusion of only Asian patients limits the generalizability of the models.

In conclusion, our study established a positive correlation between the level of pulmonary artery obstruction and both mortality and clinical deterioration. The combination of calculating nearly totally obstructed peripheral segments and sPESI provides improved predictive ability for clinical deterioration. These findings may offer physicians a tool for predicting the prognosis of patients diagnosed with acute PE.

## Supplementary material

Figure S1. Flow Diagram of the Study Cohort

Figure S2. Flow Diagram of the Validation Cohort

Table S1. Baseline Characteristics of Patients with Acute Pulmonary Embolism with and Without Clinical Deterioration in the External Validation Group

Figure S3. The Area Under the Receive Operating Characteristic for the sPESI, Miller Index, and Qanadli Index

Table S2. Univariate Regression Analysis of sPESI, Different Segments, and Models in Predicting Clinical Deterioration

Table S3. The ROC Analysis of Different Models with and without sPESI in Predicting Clinical Deterioration in the External Validation Group

Table S4. Multivariate Regression Analysis of Combination of Models Plus sPESI in Predicting Clinical Deterioration

Table S5. The NRI and IDI Comparing the Models Plus sPESI with sPESI in the External Validation Group

Table S6. The Inter- and Intra-rater Variability

Figure S4. Nomogram Combining Model 1 and sPESI

Figure S5. The Bland-Altman Plot

## Data Availability

The data that support the findings of this study are available from the corresponding author upon reasonable request.

